# A Quarter-Century of Synthetic Data in Healthcare: Unveiling Trends with Structural Topic Modeling

**DOI:** 10.1101/2025.03.30.25324911

**Authors:** Billy Ogwel, Vincent H. Mzazi, Alex O. Awuor, Gabriel Otieno, Sidney Ogolla, Bryan O. Nyawanda, Richard Omore

## Abstract

Data-driven approaches are transforming healthcare, yet acquisition of comprehensive datasets is hindered by high costs, privacy regulations, and ethical concerns. To address these challenges, synthetic data, artificially generated datasets that mimic the statistical properties of real-world data, provides a promising solution. Despite its growing adoption, the thematic landscape of synthetic data research in healthcare remains underexplored. Therefore, we applied structural topic modeling (STM) to map the research landscape of synthetic data in healthcare, revealing prevalent topics and tracking their evolution over time and across geographic locations. PubMed publications from 2000-2024 containing “synthetic data,” “artificial data,” or “simulated data” in the title/abstract were retrieved. After preprocessing the text (lowercasing, punctuation/stopword removal, stemming), structural topic modeling (STM) was performed using year and continent as covariates. The optimal number of topics (K=10) was determined using held-out likelihood and interpretability. Topic prevalence, temporal trends, and inter-topic correlations were analyzed using stacked area charts and network analysis. Analysis of 14,788 PubMed articles (2000-2024) revealed a tenfold increase in publications. Geographically, North America (48.6%) and Europe (33.5%) were primary contributors, but Asia’s share steadily rose from 2.9% to 23.1%. STM identified ten key topics, grouped into Biomedical Imaging & Signal Processing (25.2%), Synthetic Data Applications in Biomedical Research (17.7%), Computational & Statistical Methods (23.9%), and Genomics & Evolutionary Biology (33.2%) themes. We observed gradual declines in initially prominent topics including “Bayesian Modelling” (23.1% to 9.9%), “Neuroimaging” (16.0% to 9.3%), and “Image Simulation” (17.7% to 9.1%), giving ascendancy to “Synthetic Data Generation” (2.2% to 27.1%) and “Disease Modeling and Public Health” (4.8% to 11.9%) by 2024. Synthetic data research in healthcare has experienced increasing interest, marked by shifts in geographic distribution and dynamic evolution of key topics. Realizing the full potential of synthetic data requires fostering cross-disciplinary collaborations, implementing bias mitigation strategies, and establishing equitable partnerships.

**Author Summary:** In recent years, synthetic data—artificially generated datasets designed to reflect real-world information—has gained attention as a way to advance healthcare research while addressing concerns around data privacy, costs, and accessibility. Our work explores how this field has evolved over the past 25 years, identifying key research trends and shifts in geographic contributions. By analyzing over 14,000 published studies, I found that synthetic data research has grown nearly tenfold, with increasing contributions from Asia alongside traditional leaders in North America and Europe. The focus of research has also changed: earlier work emphasized medical imaging and statistical modeling, while recent studies highlight synthetic data generation and its use in disease modeling, public health, and clinical trials. Despite this progress, important gaps remain. Areas like drug discovery, mental health, and ethical considerations in artificial intelligence need further attention. By mapping these trends, our work underscores the importance of cross-disciplinary collaboration and equitable global partnerships to maximize the benefits of synthetic data in improving healthcare worldwide.

## 1. Introduction

The healthcare sector is increasingly leveraging data-driven approaches to improve patient outcomes, optimize operational efficiency, and advance medical research. However, acquiring comprehensive healthcare data is often constrained by high costs associated with advanced data acquisition techniques, privacy regulations and ethical considerations regarding patient data collection and sharing (1,2). These factors collectively impede the utilization of comprehensive data for advancing patient care and medical research. To address these challenges, synthetic data offer a promising solution.

Synthetic data, which refers to artificially generated datasets that mimic the statistical properties of real-world data without exposing sensitive information (3), mitigates data scarcity and privacy concerns, enabling a broader scope of research and experimentation without patient data exposure (4). Moreover, it facilitates the creation of diverse and representative datasets, improves AI model generalizability, and mitigates bias arising from skewed or underrepresented populations (5). [5]. Synthetic data also facilitates innovation while safeguarding patient confidentiality, making it a crucial tool for overcoming data access barriers.

While a number of reviews have examined synthetic data in healthcare (3,5–11), they predominantly focus on data generation methods (Table 1). These existing reviews of synthetic data in healthcare, while valuable, often rely on manual theme identification, introducing potential subjectivity and bias. Structural topic modeling (STM) –an advanced probabilistic unsupervised machine learning topic modeling technique designed to uncover latent themes within a collection of textual documents by incorporating document-level metadata– offers a data-driven, unbiased approach to topic extraction. Specifically, STM overcomes limitations of traditional reviews by mitigating subjective bias in theme identification, tracking the temporal evolution of synthetic data discussions, revealing shifting research focuses, and quantifying topic importance to identify research gaps and inform strategic decision-making by policymakers, funding agencies, and researchers (12,13). Therefore, the application of STM provides a complementary and distinctive perspective, beyond traditional reviews, on the existing body of review literature.

**Table 1.** Summary of review studies on synthetic data in healthcare.

| Authors | Year | Period | Topic | Database | Number of articles | Methods |
| --- | --- | --- | --- | --- | --- | --- |
| Gonzales et al. (3) | 2023 | - | Synthetic data in health care | PubMed, Scopus, and Google Scholar | 72 | A narrative review focused on thematic analysis of use cases |
| Pezoulas et al. (6) | 2024 | 2015-2024 | Synthetic data generation methods | PubMed and Scopus | 83 | Systematic review |
| Liu et al. (7) | 2025 | 2019-2023 | Deep learning approaches for synthetic data generation | Scopus, Web of Science, PubMed, and IEEE | 21 | Systematic review |
| Rujas et al. (8) | 2024 | 2014-2024 | Synthetic data generation methods: Domains, motivations, and future applications | PubMed, Scopus, and Web of Science | 42 | Scoping review |
| Goyal et al. (9) | 2024 | 2007-2024 | Synthetic Data Generation Techniques Using Generative AI | MDPI, IEEE Xplore, Science Direct, Research Gate, NeurIPS, and Arxiv | 77 | Systematic review |
| Smolyak et al. (10) | 2024 | - | Large language models and synthetic health data: progress and prospects | - | - | Synthesis of systematic scoping reviews |
| Kaabachi et al. (11) | 2025 | 2018-2024 | Privacy and utility metrics in medical synthetic data | PubMed and Embase | 24 | Scoping review |
| Murtaza et al. (5) | 2023 | - | Synthetic data generation: State of the art in health care domain | IEE, PubMed, Springer and ACM | 70 | Narrative review |

Despite its growing adoption, the thematic landscape of synthetic data research in healthcare remains underexplored. Here, we apply STM to map the research landscape of synthetic data in healthcare, revealing prevalent topics and tracking their evolution over time and across geographic locations.

## 2. Results

During the review period, 14,803 articles were published. Of these, 25 articles lacked an abstract and were excluded from further analysis that focused on 14,788 articles (Fig 1). The number of publications analyzed exhibited a fluctuating upward trajectory, increasing approximately tenfold from 138 articles in 2000 to 1,361 articles in 2024 (Fig 2).

**Fig 1.**
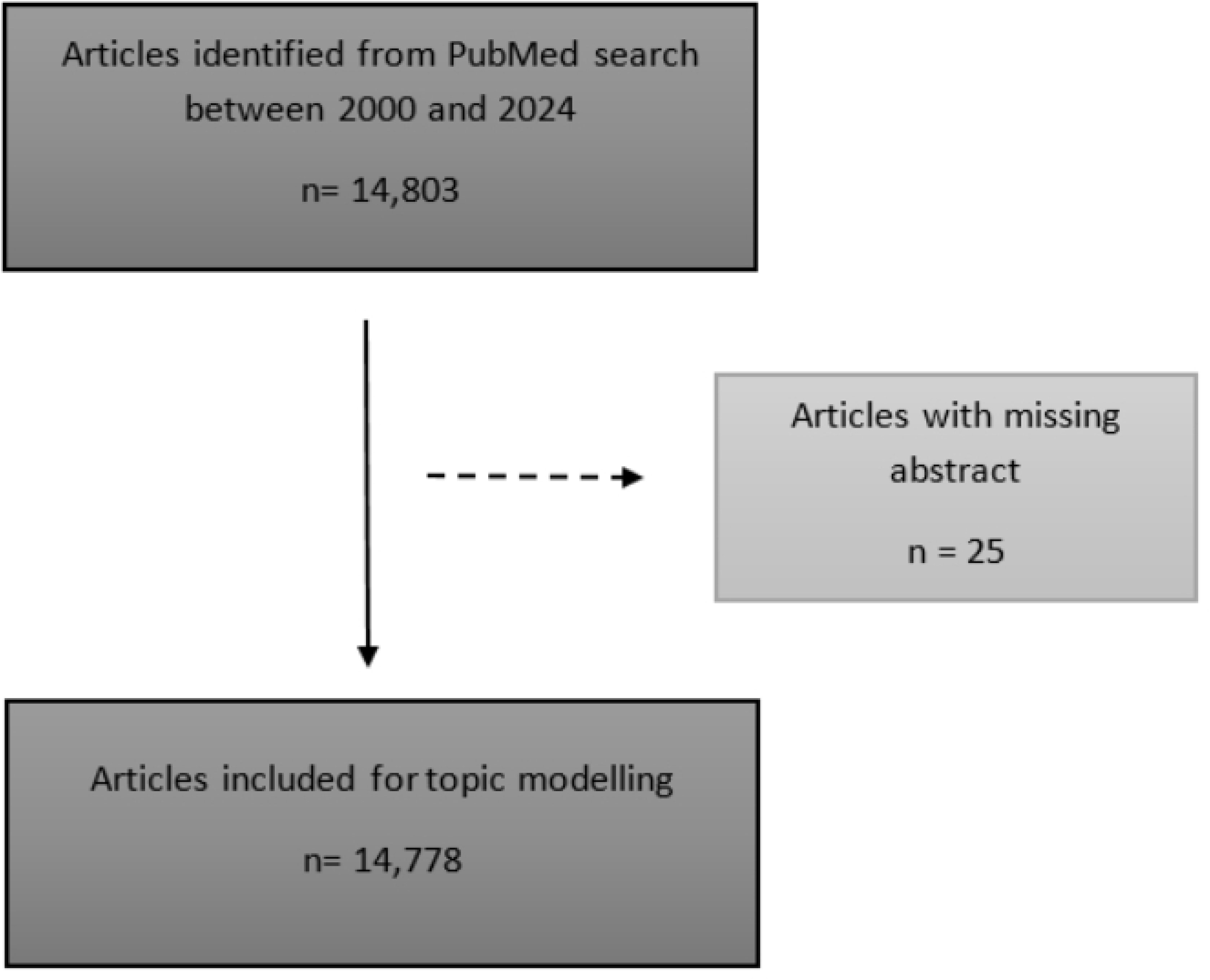
Flow diagram of included studies for synthetic data in healthcare, 2000-2024

**Fig 2.**
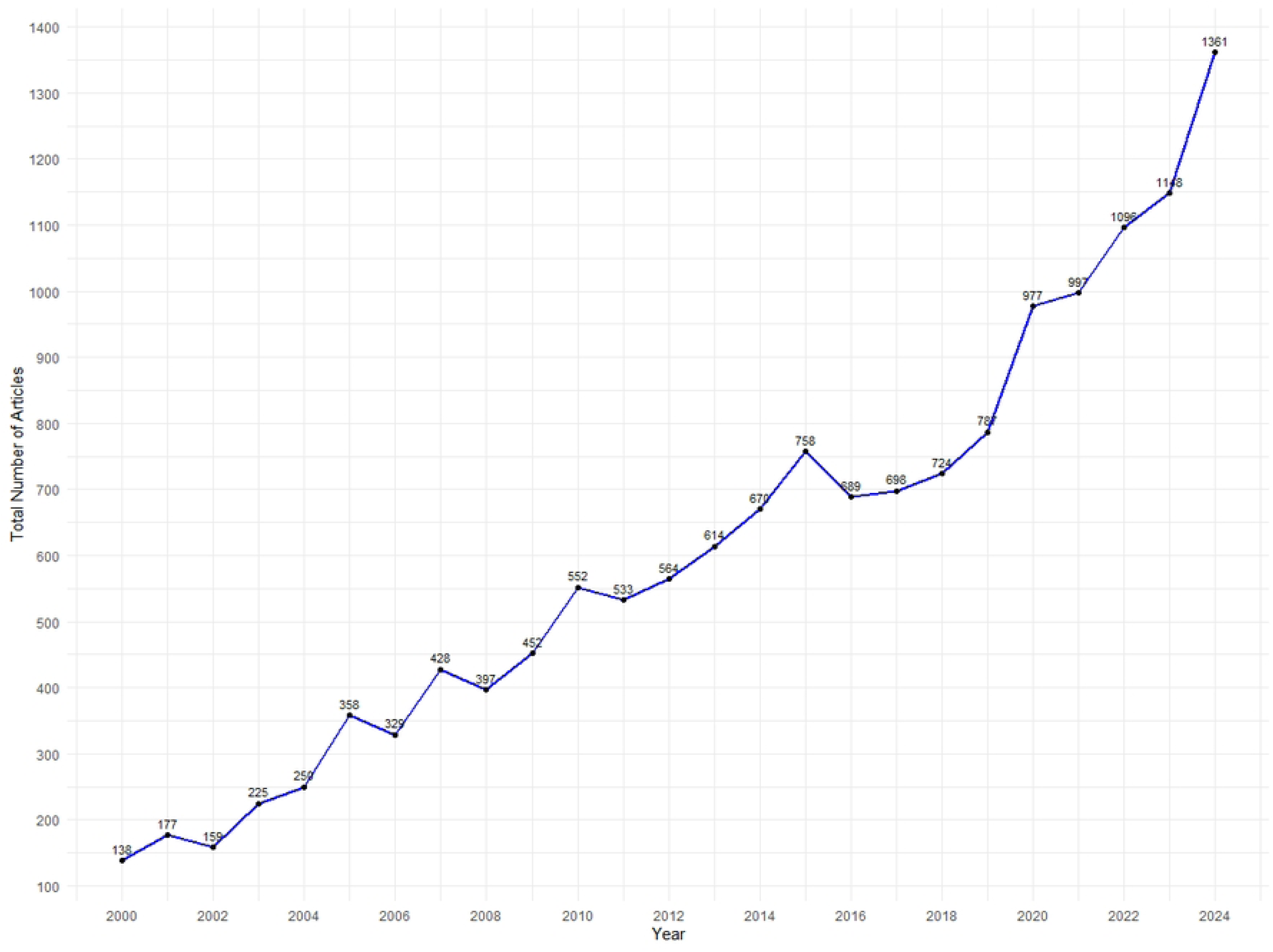
Number of articles related to synthetic data in healthcare between 2000 and 2024

### 2.1 Geographical patterns

Overall, North America (48.6%) and Europe (33.5%) were the primary contributors to the research, followed by Asia (14.4%). In contrast, Oceania (1.7%), South America (1.4%), and Africa (0.4%) had the lowest contributions. Although North America contributed the most, its share declined from 53.6% in 2000 to 41.0% in 2024. A similar trend was observed for Europe, decreasing from 39.9% to 32.3% over the same period. Conversely, Asia experienced a steady rise, increasing from 2.9% in 2000 to 23.1% in 2024. Oceania and South America demonstrated minor fluctuations, ranging from 2.9% to 1.8% and 0.7% to 1.2%, respectively, between 2000 and 2024. Africa’s contribution remained low but relatively stable, starting at 0.6% in 2006 and ending at 0.4% in 2024 (Fig 3). At the country level, the top five contributors to the research were the United States of America (6,897 articles), China (1,066), the United Kingdom (1,051), Germany (966), and France (622).

**Fig 3.**
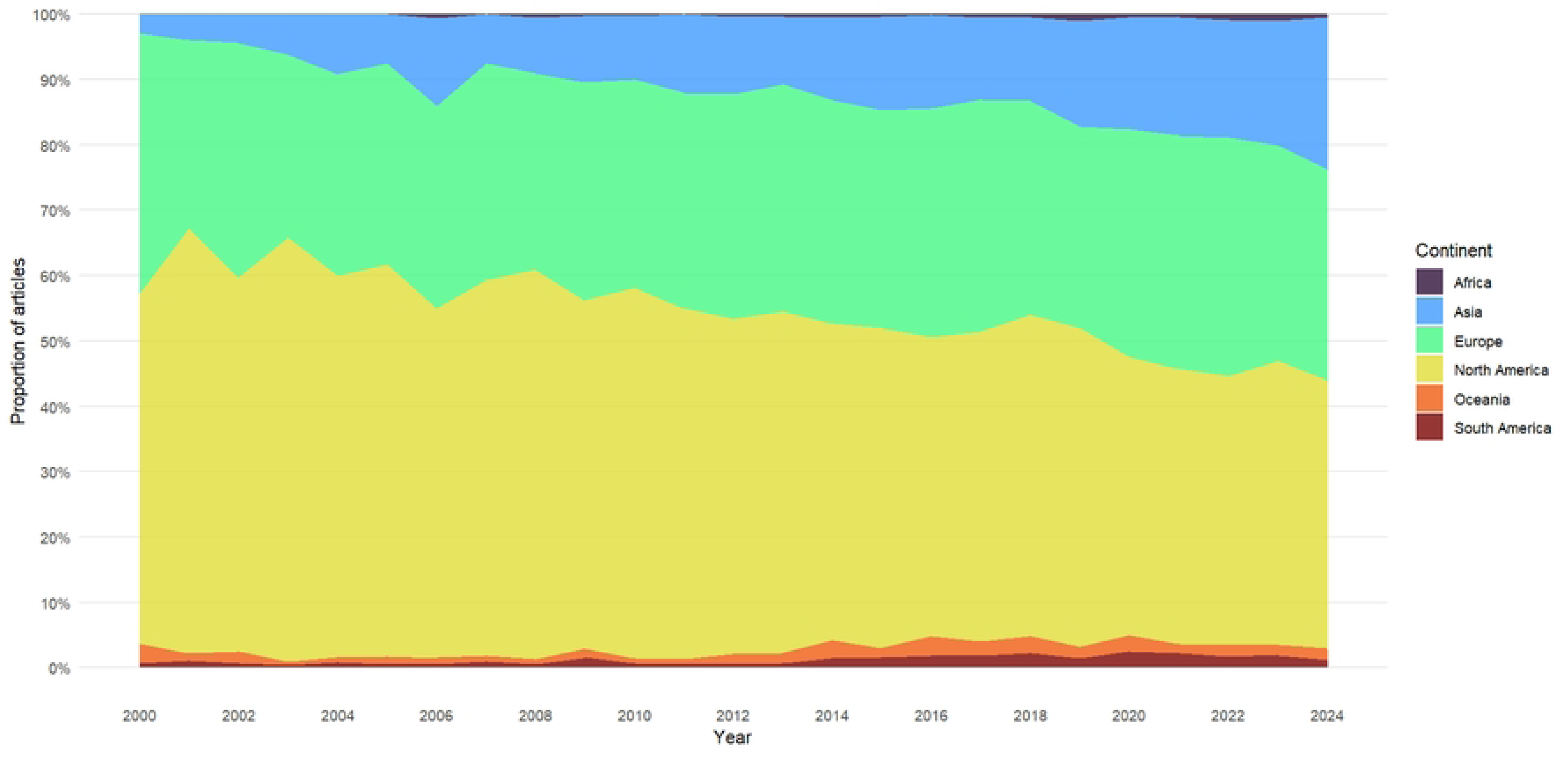
Annual proportion of synthetic data research in healthcare by continent (2000-2024)

### 2.2 Topic identification and prevalence

Table 2 presents the identified topics along with key terms for each metric, as well as the thematic areas encompassing the ten topics. Fig 4 presents word clouds visualizing the most frequent words for each topic, ranked by the FREX criterion. Word size corresponds to word frequency. Among these, Topic 8: Bayesian Modeling & Inference (16.2%), Topic 1: Neuroimaging & Signal Processing (13.8%), Topic 5: Clinical Trials and Statistical Inference (11.8%), and Topic 7: Image Simulation and Physical Modeling (11.5%) emerged as the most prominent topics in synthetic data research in healthcare. In 2000, the most important topics were Topic 8: Bayesian Modeling & Inference (23.1%), Topic 7: Image Simulation and Physical Modeling (17.7%), and Topic 1: Neuroimaging & Signal Processing (16.0%). By 2024, the focus shifted to Topic 4: Synthetic Data Generation (27.1%), Topic 2: Disease Modeling and Public Health (11.9%), and Topic 5: Clinical Trials and Statistical Inference (9.9%), reflecting an evolving research landscape (Fig 5).

**Fig 4.**
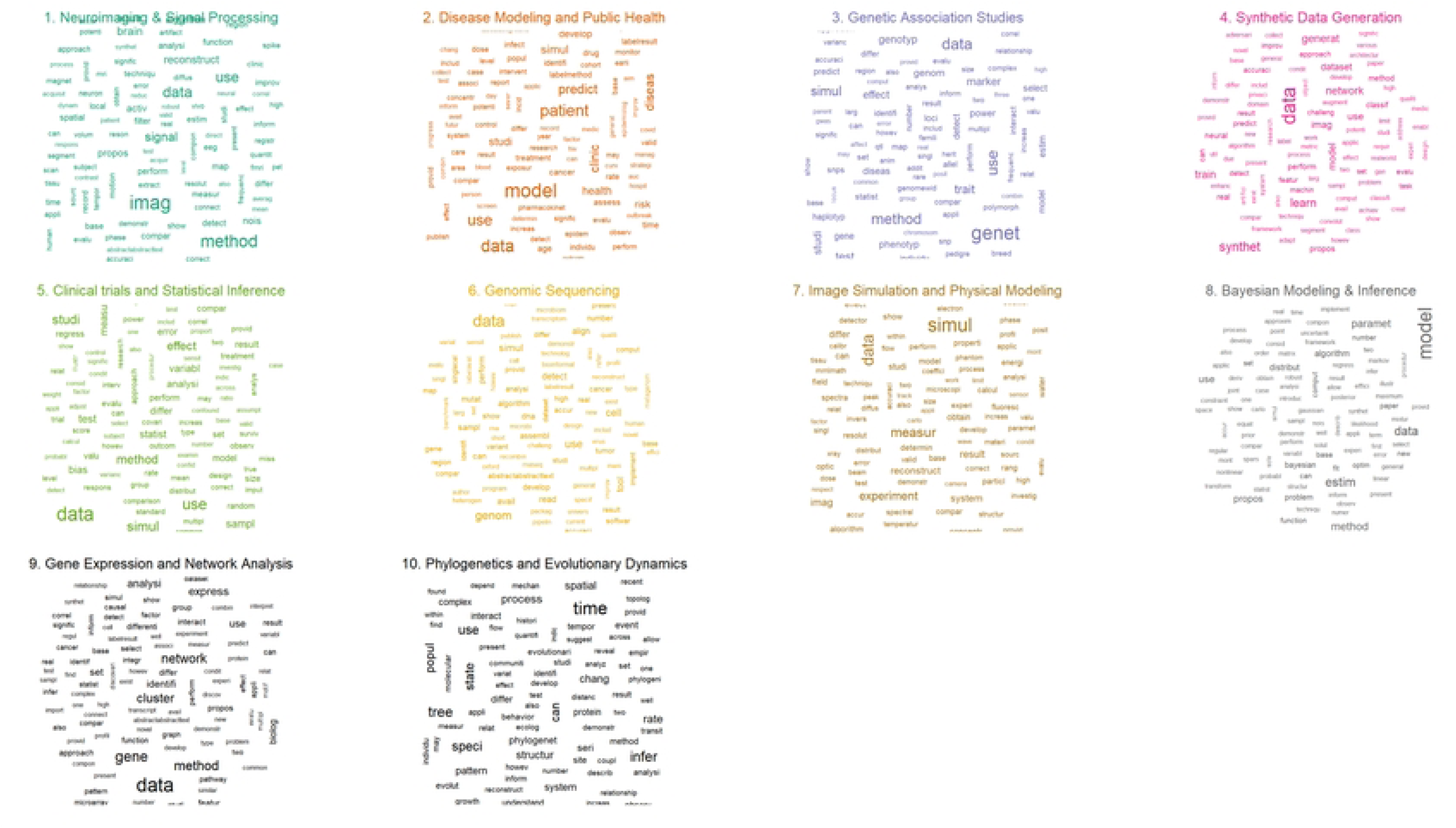
Word cloud figure for the ten topics in synthetic data research in healthcare, 2000-2024

**Fig 5.**
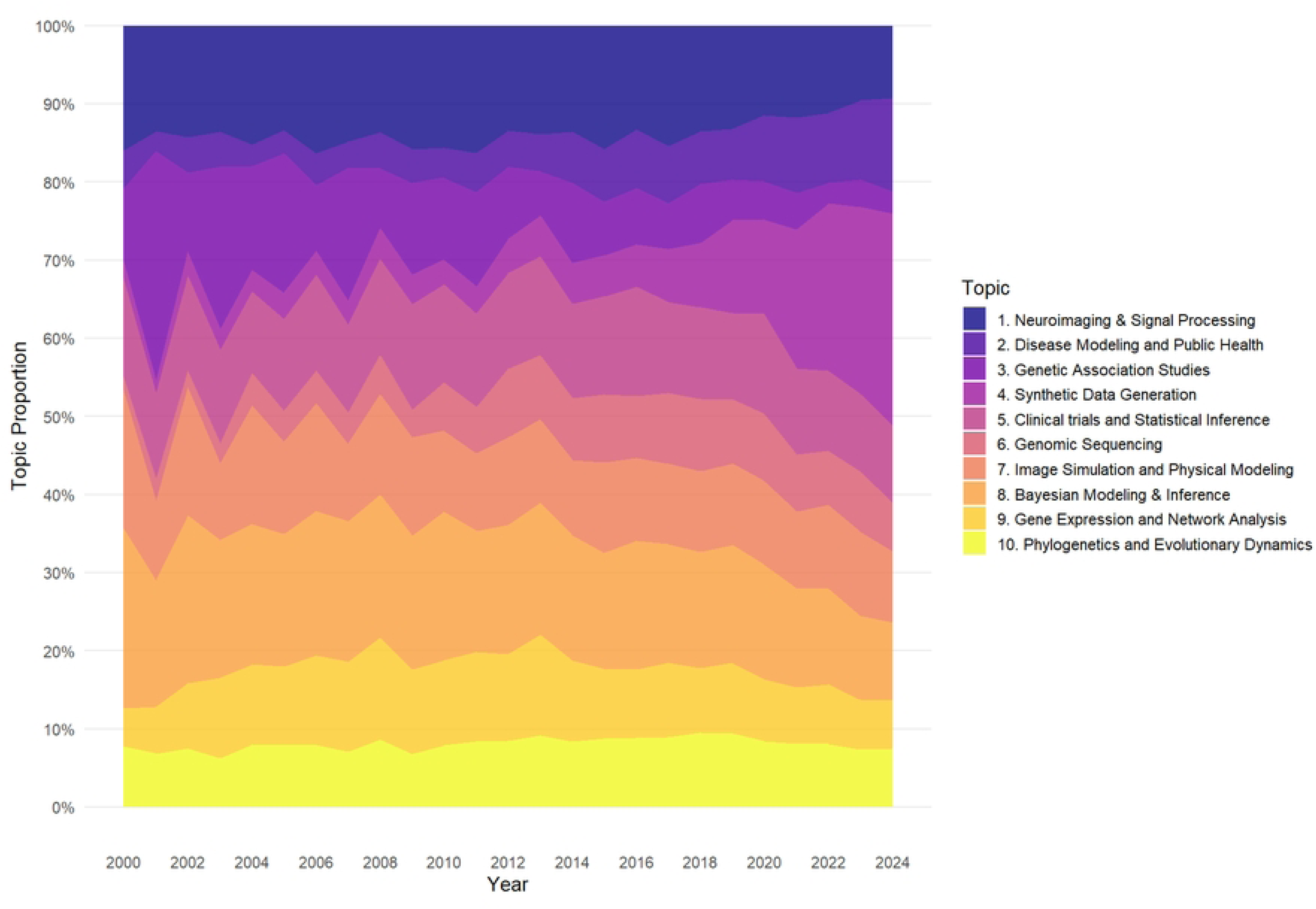
Temporal patterns in synthetic data research in healthcare topics, 2002-2024

**Table 2.**
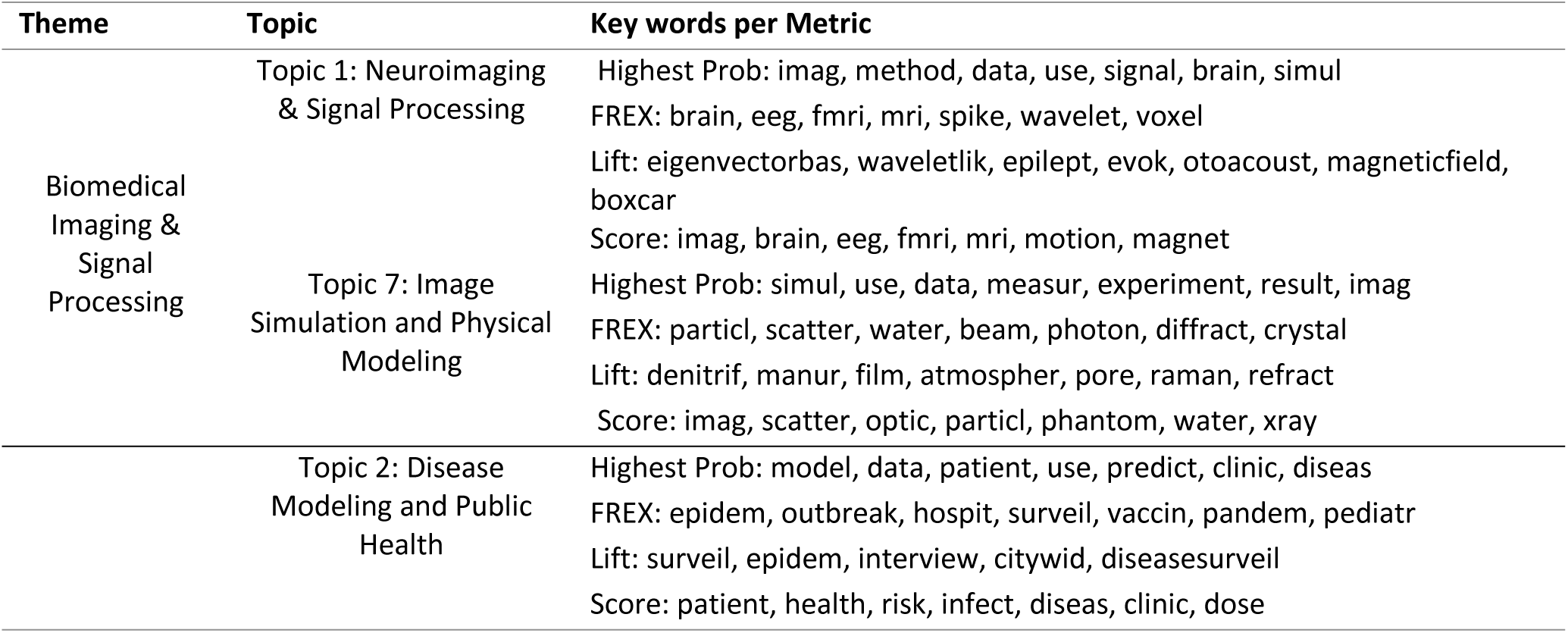
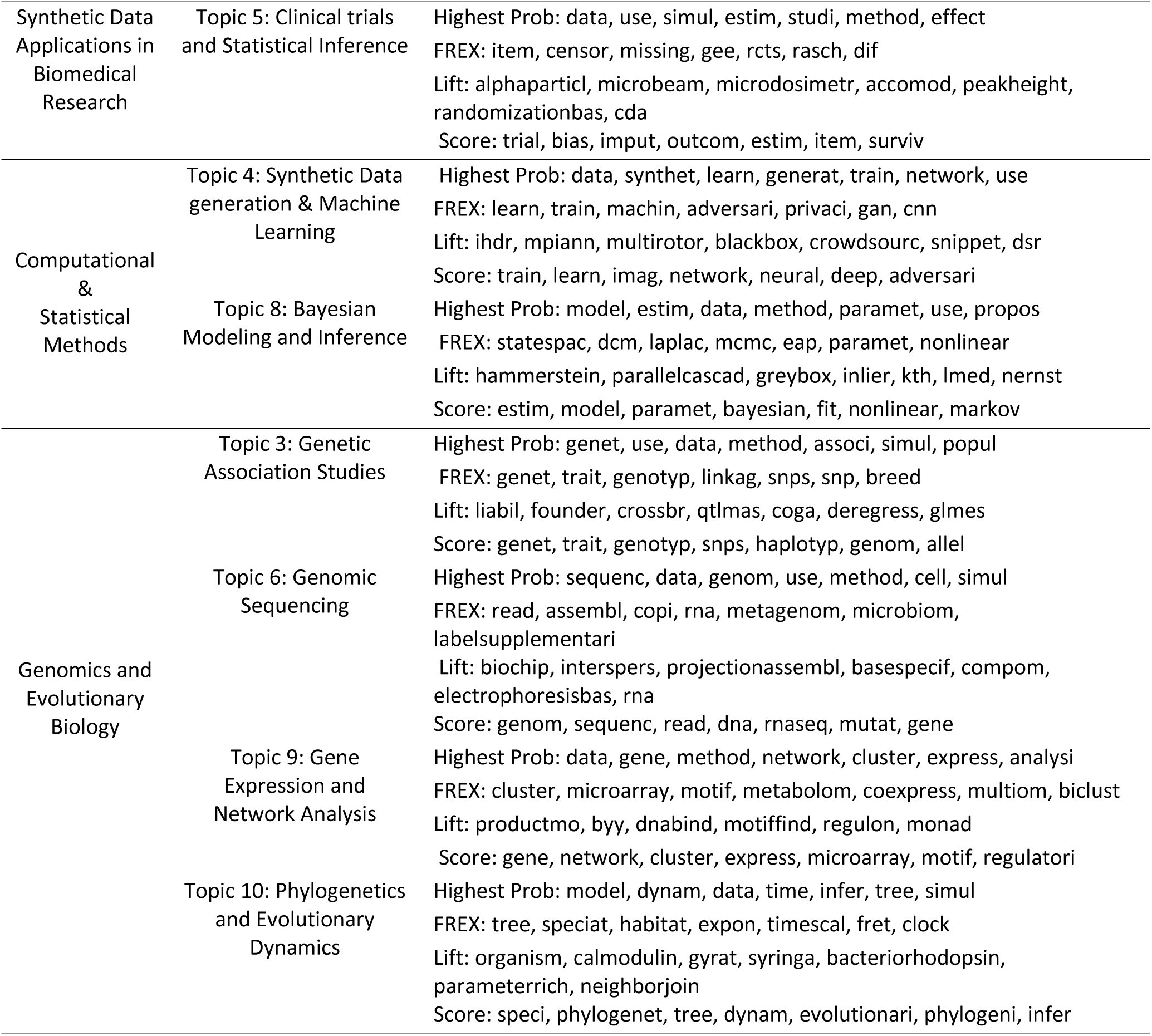
Themes, topic labels, and key terms in synthetic data research for healthcare.

### 2.3 Thematic groups and temporal dynamics

The identified topics were grouped into four thematic areas:

#### 2.3.1 Theme 1: Biomedical Imaging & Signal Processing

This theme encompasses two topics: Topic 1: Neuroimaging & Signal Processing, and Topic 7: Image Simulation and Physical Modeling (Table 2), collectively representing 25.2% of all evaluated publications. Topic 1 accounted for 13.8%, while Topic 7 contributed 11.4%. Over time, both topics exhibited a downward trend, with Topic 1 declining from 16.0% to 9.3% and Topic 7 decreasing from 17.7% to 9.1%, between 2000 and 2024 (Fig 5).

#### 2.3.2 Theme 2: Synthetic Data Applications in Biomedical Research

Comprising Topic 2 (Disease Modeling and Public Health) and Topic 5 (Clinical Trials and Statistical Inference) (Table 2), the Synthetic Data Applications in Biomedical Research theme (Table 2) represented 17.7% of the total publications. Topic 2 and Topic 5 individually contributed 5.9% and 11.8%, respectively. These topics had differing temporal trajectories: Topic 2 saw growth from 4.8% in 2000 to 11.9% in 2024, whereas Topic 5 exhibited a marginal decrease from 12.6% to 9.9% during the same timeframe (Fig 5).

#### 2.3.3 Theme 3: Computational & Statistical Methods

Comprising Topics 4 (Synthetic Data Generation) and 8 (Bayesian Modeling & Inference) (Table 2), the Computational & Statistical Methods theme accounted for 23.9% of the research output. With individual contributions of 7.6% and 16.2% respectively, these topics exhibited contrasting temporal patterns. While Topic 4 rose dramatically, from 2.2% in 2000 to become the leading topic in 2024, representing 27.1% of research topics, topic 8 declined (23.1% to 9.9%), (Fig 5).

#### 2.3.4 Theme 4: Genomics & Evolutionary Biology

Comprising four diverse topics – 3 (Genetic Association Studies), 6 (Genomic Sequencing), 9 (Gene Expression and Network Analysis), and 10 (Phylogenetics and Evolutionary Dynamics) – the Genomics and Evolutionary Biology theme (Table 2) accounted for a significant 33.2% of the analyzed research. The temporal trends within this theme were varied: Topic 3 experienced a decline (9.1% to 2.9%), Topic 6, and Topic 9 exhibited upward trends (1.7% to 6.1%, and 4.9% to 6.2%, respectively), and Topic 10 remained relatively stable (7.7% to 7.4%) over the study period (Fig 5).

### 2.4 Topic co-occurrence and correlations

The interconnections between different research topics in synthetic data research in healthcare are shown in the topic network diagram (Fig 6). The strongest correlations were between “Bayesian modelling and inference” (Topic 8) and “Synthetic Data Generation” (Topic 4) indicating a significant role of probabilistic modeling in data generation techniques. Additionally, “Neuroimaging & Signal Processing” (Topic 1) and “Clinical Trials & Statistical Inference” (Topic 5) had a strong correlation suggesting that neuroimaging applications heavily rely on statistical inference for clinical research and trial evaluations. “Genetic Association Studies” (Topic 3) and “Genomic Sequencing” (Topic 6) were also strongly connected, reflecting the inherent relationship between genome-wide sequencing and association studies in genetic research. (Fig 5). Beyond these specific connections, certain topics acted as key intermediaries, bridging different research areas. “Synthetic Data Generation” (Topic 4) served as a bridge between “Bayesian Inference” (Topic 8) and “genomic studies” (Topic 6), highlighting its role in linking statistical modeling with data generation for genomics. “Neuroimaging & Signal Processing” (Topic 1) connected biomedical imaging techniques with statistical inference, indicating its involvement in both data acquisition and analytical frameworks. Finally, “Disease Modeling & Public Health” (Topic 2) linked multiple topics, reflecting its interdisciplinary nature and integration of statistics, genetics, and computational modeling approaches.

**Fig 6.**
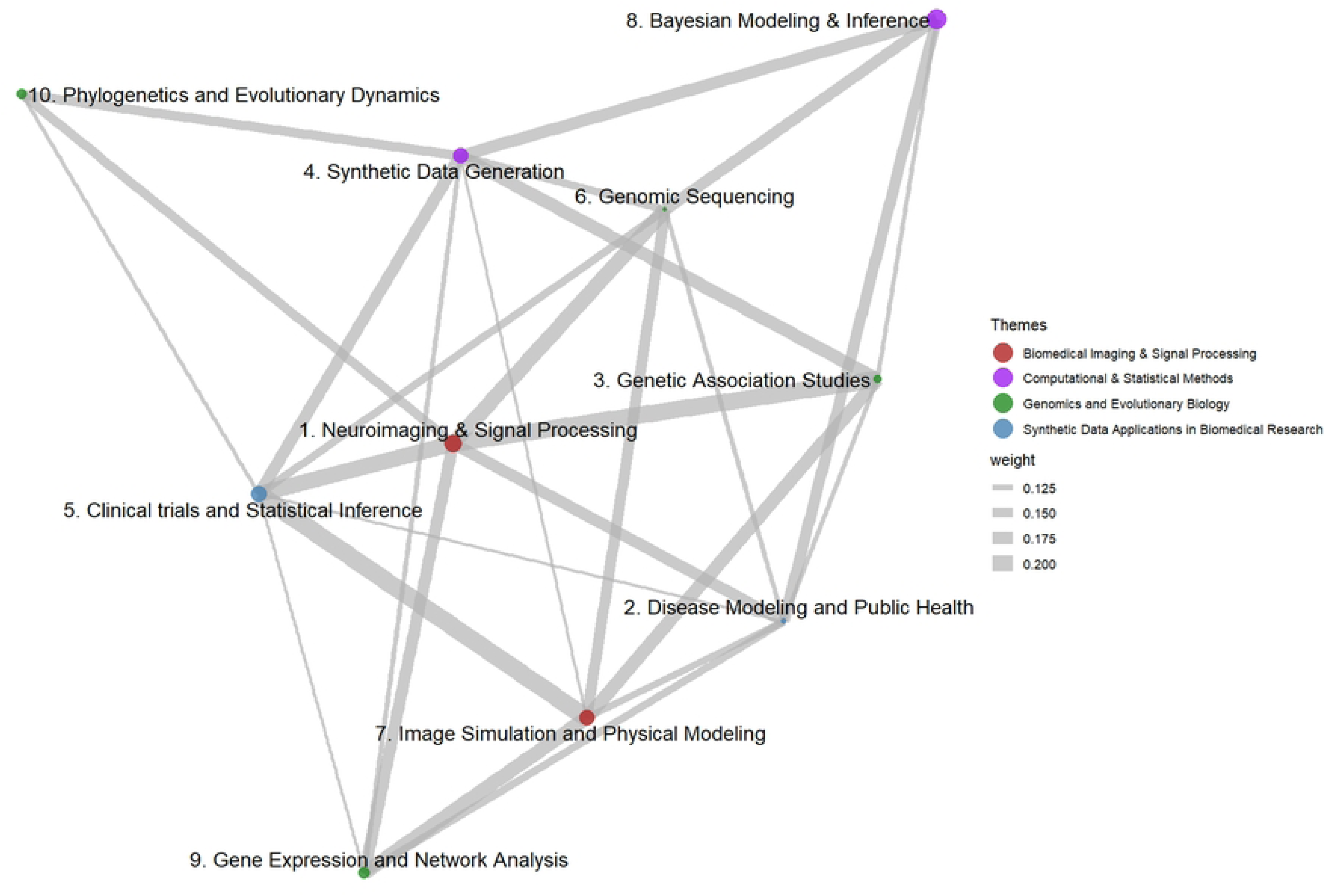
Topic network in synthetic data research in healthcare, 2000-2024.

## 3. Discussion

We mapped 25 years of synthetic data research in healthcare using a structural topic modelling approach. The research output grew nearly tenfold, from 138 articles in 2000 to 1,361 in 2024, reflecting a rising interest in synthetic data applications in healthcare. Furthermore, we observed substantial changes in the geographic distribution of research activity and a dynamic evolution of key research topics. Specifically, initially prominent topics including “Bayesian Modelling” (23.1% to 9.9%), “Neuroimaging” (16.0% to 9.3%), and “Image Simulation” (17.7% to 9.1%) gradually decreased as the research focus shifted to “Synthetic Data Generation” (27.1%), “Disease Modeling and Public Health” (11.9%), and “Clinical Trials and Statistical Inference” (9.9%) in 2024.

### 3.1 Geographical landscape

Geographically, North America and Europe were initially the dominant contributors, but Asia’s contribution significantly increased over time. Overall, Oceania, South America, and Africa contributed minimally (<2% each). The observed geographic patterns reflect both regional dominance and disparity in synthetic data research in healthcare. North America and Europe’s historical leadership stems from established institutions, robust funding, strong industry-academia collaborations, and mature regulatory frameworks. Conversely, Asia’s rising contribution, particularly from China driven largely by state-led investment in research (2.4% of GDP in 2022) prioritizing AI and healthcare, signals a shifting research landscape (22). The persistent Global North-South disparity (23–25), exacerbated by limited funding, infrastructure deficits, brain drain, and weak regulatory frameworks in Africa, South America, and Southeast Asia, necessitates capacity-building investments and equitable North-South partnerships to foster a more balanced global research eco-system.

### 3.2 Thematic landscape and temporal dynamics

The thematic contributions and temporal dynamics in synthetic data research reflect evolving technological priorities, healthcare demands, and methodological advancements. Biomedical Imaging & Signal Processing theme (25.2%), once dominant, has seen gradual declines in subtopics like “Neuroimaging” (16.0% to 9.3%) and “Image Simulation” (17.7% to 9.1%). This trend may stem from the maturation of imaging technologies, where foundational innovations in computed tomography (CT), magnetic resonance imaging (MRI), and signal processing have already been integrated into clinical workflows, reducing the urgency for novel breakthroughs (26,27). Additionally, the rise of computationally intensive methods in other areas, such as synthetic data, may have diverted the focus from these areas.

The Computational & Statistical Methods theme (23.9%) illustrates a pivotal shift: “Synthetic Data Generation” surged from 2.2% to 27.1%, overshadowing declines in “Bayesian Modeling & Inference” (23.1% to 9.9%). This shows the growing demand for scalable, AI-driven solutions in research and healthcare, where synthetic data addresses challenges like data privacy, scarcity of labeled datasets, and the need to train robust machine learning models (4). Bayesian methods, while robust, may have waned due to their computational complexity and the preference for faster, data-driven approaches in large-scale applications (28). The Synthetic Data Applications in Biomedical Research theme (17.7%) highlights contrasting trajectories: “Disease Modeling & Public Health” grew (4.8% to 11.9%), likely driven by global crises like the COVID-19 pandemic, which underscored the need for predictive modelling to inform policy and outbreak management (29). Conversely, “Clinical Trials & Statistical Inference” declined (12.6% to 9.9%), possibly due to increasing regulatory scrutiny and the difficulty of translating synthetic data into accepted clinical endpoints.

Genomics & Evolutionary Biology (33.2%), the largest theme, reveals subfield variability. Synthetic genomic data addresses data scarcity and privacy concerns by enabling researchers to share datasets mimicking real sequences without exposing individual identities, facilitating studies of rare variants and diseases, modeling evolutionary dynamics, and validating algorithms (30–33). It supports hypothesis testing via evolutionary simulations, allows for controlled benchmarking and bias mitigation, and facilitates large-scale population genomics and high-risk climate adaptation modeling. Furthermore, synthetic data democratizes genomic research by providing usable datasets for resource-limited settings and addressing ethical concerns in indigenous communities, promoting equitable access and collaboration in the field. While synthetic genomic data is increasingly utilized, concerns exist regarding its fidelity and ability to capture complex genomic phenomena such as epistasis and epigenetic regulation (34,35). Additionally, establishing statistical validation standards is crucial to ensure the trustworthiness of synthetic genomic datasets. Temporally, “Genomic Sequencing” (1.7% to 6.1%) and “Gene Expression Analysis” (4.9% to 6.2%) grew—fueled by reducing sequencing costs and the rise of precision medicine—, while “ Genetic Association Studies” declined (9.1% to 2.9%), potentially due to saturation in identifying common genetic variants and shifting focus to functional genomics (36).

### 3.3 Topic co-occurrence and correlations

The observed interconnections in the topic network—between “Bayesian Modeling” and “Synthetic Data Generation”, “Neuroimaging and Clinical Trials”, and “Genetic Association Studies” and “Genomic Sequencing”—stem from methodological synergies and evolving research demands. “Bayesian Modeling underpins “Synthetic Data Generation” by providing probabilistic frameworks to simulate realistic datasets while preserving privacy, critical for genomics and clinical applications. Neuroimaging’s link to “Clinical Trials” arises from the need for synthetic biomarkers to optimize trial designs and statistical tools to analyze complex imaging data, particularly for neurological disorders. “Genetic Association Studies” and “Genomic Sequencing” are inherently interdependent, as sequencing generates the raw data for identifying genotype-phenotype relationships, with synthetic data enabling scalable studies on rare variants or diverse populations (37). The bridging roles of “Synthetic Data Generation”, “Neuroimaging”, and “Disease Modeling” reflect their cross-disciplinary utility. Synthetic Data connects computational methods to applied fields by enabling algorithm validation and scalable simulations. Neuroimaging bridges biomedical engineering (data acquisition) and clinical research (diagnostic AI tools), while Disease Modeling integrates genomics, epidemiology, and public health to address crises like pandemics. These linkages are driven by technological convergence, demands for reproducibility, and global challenges requiring holistic approaches.

### 3.4 Research gaps

Despite the diversity of themes identified in this analysis of synthetic healthcare data, key gaps remain. Drug discovery, synthetic Electronic Health Records (EHRs), and personalized medicine beyond genomics are underrepresented due to challenges in molecular modeling, regulatory barriers, fragmented standards, and data scarcity. Emerging areas like telemedicine and mental health face similar limitations related to data complexity and ethical concerns. Furthermore, health economics and ethical AI applications require more explicit attention to bias mitigation. These gaps stem from technical limitations in generative modeling, ethical considerations, interdisciplinary silos, and nascent development. Addressing these challenges demands cross-disciplinary collaborations, specialized tools for complex data types, proactive fairness auditing, and engagement with evolving regulatory landscapes to maximize the impact of synthetic data across healthcare.

### 3.5 Limitations

This study’s methodology is subject to some limitations. The search strategy, restricted to specific keywords in titles/abstracts and reliance on PubMed, introduces potential selection bias by excluding relevant literature using alternative terminology or indexed in other databases. Geographic classification based solely on first-author affiliation may misrepresent multinational collaborations and regional contributions. While language bias was minimized via English titles/abstracts, exclusion of non-English publications is still possible. Future analyses should mitigate these issues through expanded search terms and data sources, expert validation, and refined geographic classification methods to enhance robustness and representativeness.

### 3.6 Conclusion

Through structural topic modeling analysis of 25 years of synthetic data research in healthcare this study reveals significant growth and evolution of topics. Research output has increased nearly tenfold, accompanied by a shift in geographic contributions, with Asia’s presence growing alongside the historical dominance of North America and Europe, although a persistent Global North-South disparity remains. Thematic foci have evolved, with increased emphasis on “Synthetic Data Generation”, “Disease Modeling and Public Health” and “Clinical Trials and Statistical Inference”. Methodological interconnections are evident. Despite these advances, notable gaps exist in areas such as drug discovery, synthetic EHRs, personalized medicine beyond genomics, telemedicine, mental health applications, health economics, and ethical AI, highlighting the need for cross-disciplinary collaborations, bias mitigation strategies, and equitable partnerships to fully realize the potential of synthetic data in healthcare.

## 4. Materials and Methods

### 4.1 Data Collection

We systematically retrieved relevant publications from PubMed using a search strategy that targeted articles containing terms related to synthetic data in the title or abstract: (’“Synthetic data” OR “Artificial data” OR “Simulated data”). The batch_pubmed_download function was used to systematically retrieve articles in XML format while adhering to PubMed API rate limits (14). We included articles published between 2000 and 2024 with an abstract.

### 4.2 Data Processing

Each downloaded XML file was assigned a unique timestamp-based filename to prevent duplication. The downloaded XML files were processed to extract key bibliographic information, including title, abstract, publication year and first author affiliations. Metadata variables were extracted, including publication year and country. Countries were categorized into continents (Africa, Asia, Europe, North America, South America, and Oceania). To ensure data integrity, articles were assessed for missing values in key fields, duplicate titles and extracted fields were cleaned to remove HTML/XML artifacts.

### 4.3 Text Preprocessing

Titles and abstracts from the dataset were combined to form a text corpus. The corpus was preprocessed using the tm package in R in a process that involved conversion of text to lowercase, removal of punctuation, numbers and English stopwords and stripping of extra whitespace (15). Thereafter we stemmed all words using the Porter stemming algorithm (16). A Document-Term Matrix (DTM) was then created, transformed into a matrix format, and transposed to generate a term matrix.

### 4.4 Topic Modelling

Structural Topic Modeling is an advanced probabilistic unsupervised machine learning topic modeling technique designed to uncover latent themes within a collection of textual documents by incorporating document-level metadata (17–19). Preprocessed text data was formatted for STM analysis. Documents were structured into a list format compatible with the stm package, and the vocabulary was extracted from the term matrix.

To determine the optimal number of topics, a search was performed using searchK(), which evaluates model fit using held-out likelihood (15). A range of topic numbers (K = 5, 10, 15, 20, 25, 30) was tested, incorporating year and continent as prevalence covariates. The optimal number of topics (K) was selected by identifying the “elbow point” in the likelihood plot, balancing semantic coherence and exclusivity, and evaluating the interpretability of the resulting topics (20) (Fig 7). A final STM model was fitted with K = 10 topics, using year and continent as prevalence covariates. The Expectation-Maximization (EM) algorithm was run for 150 iterations, initializing with a Latent Dirichlet Allocation (LDA) approach (21).

**Fig 7.**
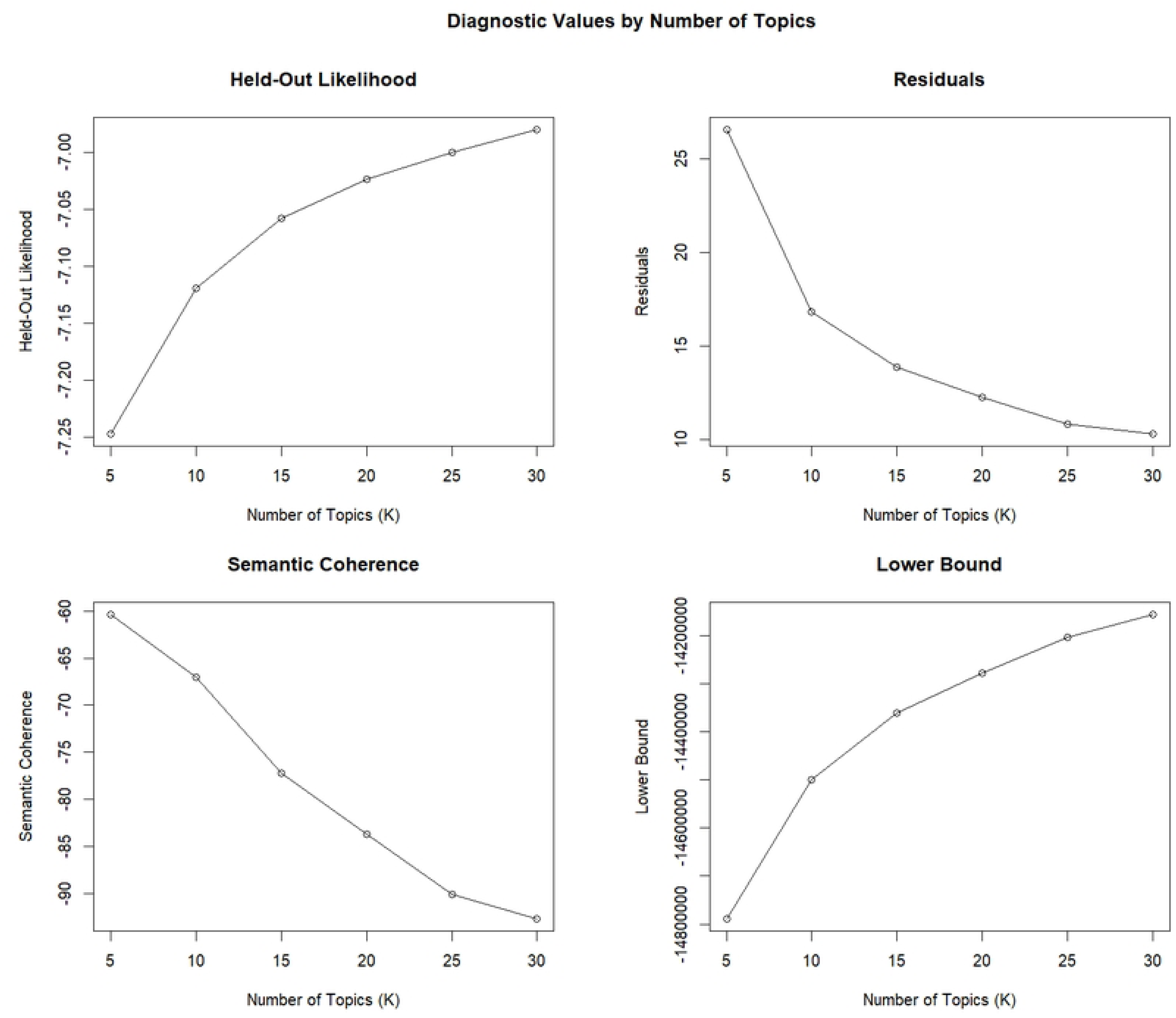
Diagnostic values by the number of topics for synthetic data in healthcare, 2000-2024

The top terms associated with each topic were extracted using the labelTopics() function. Based on interpretability, topics were assigned descriptive labels reflecting their thematic content. Furthermore, to enhance the interpretability of the topics generated by the STM model, word clouds were created for each topic, highlighting the most frequently associated words (21). The cloud() function from the stm package was used, with word sizes scaled to reflect their relative importance within each topic. To enhance interpretability, topics were then grouped into broader thematic categories.

To assess the temporal trends in STM research, we computed the annual count of articles for each continent and standardized the data to ensure a complete set of year-continent combinations. Missing values were replaced with zero to maintain data consistency. We then calculated the annual proportion of articles per continent to facilitate a comparative analysis. We employed 100% stacked area chart to illustrate the temporal distribution of STM research by continent. To examine how research topics evolved over time, we linked topic prevalence data with publication years. We generated 100% stacked area chart to depict changes in topic proportions across years.

We assessed relationships between topics using correlation-based network analysis (15,21). A topic correlation matrix was derived from the STM model, with edges retained for correlations above a predefined threshold (0.1). An adjacency matrix was created to construct a graph-based network, which was visualized using the igraph and ggraph packages. To improve readability, we, scaled node sizes based on topic prevalence, colored nodes according to their thematic classification, and weighted edges based on correlation strength. Additionally, a force-directed layout was employed to enhance network interpretability, and labels were added to identify key topics.

Descriptive analysis using percentages and topic modeling were performed using R version 4.4.1 (R Foundation for Statistical Computing, Vienna, Austria).

### 4.5 Ethical approval

Ethical approval was not required for this study as it did not involve human or animal participants.

## Declarations

### Acknowledgements

Not Applicable

### Consent for publication

Not Applicable

### Protocol and Registration

A study-specific protocol was not developed for this work, nor was the protocol registered.

### Availability of data and materials

The programming code for R is available on GitHub: https://github.com/bogwel/Synthetic_data_STM/tree/main

### Author Contributions

BO conceived the study, BO, VM, BON, AOA, GO and RO contributed to study design and implementation. BO and BON analyzed and interpreted the data. BO drafted the manuscript and all authors critically reviewed the manuscript for intellectual content and approved the final manuscript.

## Data Availability

We retrieved publicly available metadata from PubMed using a structured search for "synthetic data," "artificial data," and "simulated data" in titles and abstracts. The search strategy is detailed in the Methods section.

